# Dimethyl fumarate in patients admitted to hospital with COVID-19 (RECOVERY): a randomised, controlled, open-label, platform trial

**DOI:** 10.1101/2022.09.23.22280285

**Authors:** RECOVERY Collaborative Group, Peter W Horby, Leon Peto, Natalie Staplin, Mark Campbell, Guilherme Pessoa-Amorim, Marion Mafham, Jonathan R Emberson, Richard Stewart, Benjamin Prudon, Alison Uriel, Christopher A Green, Devesh J Dhasmana, Flora Malein, Jaydip Majumdar, Paul Collini, Jack Shurmer, Bryan Yates, J Kenneth Baillie, Maya H Buch, Jeremy N Day, Saul N Faust, Thomas Jaki, Katie Jeffery, Edmund Juszczak, Marian Knight, Wei Shen Lim, Alan Montgomery, Andrew Mumford, Kathryn Rowan, Guy Thwaites, Richard Haynes, Martin Landray

## Abstract

**Background:** Dimethyl fumarate (DMF) is an anti-inflammatory drug that has been proposed as a treatment for patients hospitalised with COVID-19

**Methods:** This randomised, controlled, open-label platform trial (Randomised Evaluation of COVID-19 Therapy [RECOVERY]), is assessing multiple possible treatments in patients hospitalised for COVID-19. In this initial assessment of DMF, performed at 27 UK hospitals, eligible and consenting adults were randomly allocated (1:1) to either usual standard of care alone or usual standard of care plus DMF 120mg twice daily for 2 days followed by 240mg twice daily for 8 days, or until discharge if sooner. The primary outcome was clinical status on day 5 measured on a seven-point ordinal scale, assessed using a proportional odds model. Secondary outcomes were time to sustained improvement in clinical status, time to discharge, day 5 peripheral blood oxygenation, day 5 C-reactive protein, and improvement in day 10 clinical status. The trial is registered with ISRCTN (50189673) and clinicaltrials.gov (NCT04381936).

**Findings:** Between 2 March 2021 and 18 November 2021, 713 patients were enrolled in the DMF evaluation, of whom 356 were randomly allocated to receive usual care plus DMF, and 357 to usual care alone. 95% of patients were receiving corticosteroids as part of routine care. There was no evidence of a beneficial effect of DMF on clinical status at day 5 (common odds ratio of unfavourable outcome 1.12; 95% CI 0.85-1.46; p=0.42). There was no significant effect of DMF on any secondary outcome. As expected, DMF caused flushing and gastrointestinal symptoms, each in around 6% of patients, but no new adverse effects were identified.

**Interpretation:** In adults hospitalised with COVID-19, DMF was not associated with an improvement in clinical outcomes.

**Funding:** UK Research and Innovation (Medical Research Council) and National Institute of Health Research (Grant ref: MC_PC_19056).

## INTRODUCTION

Severe COVID-19 is characterised by marked inflammation of the lungs, which causes respiratory failure and is usually associated with elevated circulating inflammatory markers such as C-reactive protein (CRP) and interleukin-6 (IL-6).^1–4^ This has led to the evaluation of several different kinds of immunomodulation in the treatment of severe COVID-19. Corticosteroids, IL-6 inhibitors, and Janus kinase inhibitors have all been found to reduce mortality in hospitalised patients, although the risk of death remains high even when these treatments are used.^5–8^ The effectiveness of these drugs proves that inflammation is a modifiable cause of death in patients with COVID-19, and suggests that other ways of modifying the immune response might also be beneficial.

Inflammasomes are part of the innate immune response, and have been proposed as important mediators of COVID-19 lung disease.^9,10^ These cytosolic pattern recognition receptor systems stimulate the release of proinflammatory cytokines and activate inflammatory cell death (pyroptosis).^11^ In COVID-19, the degree of inflammasome activation, particularly of the NLRP3 inflammasome, correlates with disease severity.^12^ However, although this pathway has been identified as a promising therapeutic target, treatment with colchicine, which inhibits NLRP3 inflammasome activation, does not improve outcomes in hospitalised patients.^13^ Dimethyl fumarate (DMF) is thought to inhibit NLRP3 inflammasome activation via a different mechanism to colchicine, by inactivating gasdermin D, and has been found to have anti-viral and anti-inflammatory effects against SARS-CoV-2 *in vitro*^14,15^. It is licensed to treat relapsing remitting multiple sclerosis and plaque psoriasis, and is generally well-tolerated, although often associated with flushing and gastrointestinal symptoms on initiation^16,17^. As part of the UK COVID-19 Therapeutics Advisory Panel (CTAP) review of possible therapeutics for evaluation in clinical trials, CTAP recommended to the RECOVERY chief investigators that DMF be investigated in an early phase assessment among hospitalised patients, with subsequent assessment in a larger trial of its effect on mortality if there was evidence of efficacy on surrogate outcomes. Here we report the results of an early phase randomised assessment of DMF in patients hospitalised with COVID-19, performed as part of the RECOVERY platform trial.

## METHODS

### Study design and participants

The Randomised Evaluation of COVID-19 therapy (RECOVERY) trial is an investigator-initiated, streamlined, individually randomised, controlled, open-label, platform trial to evaluate the effects of potential treatments in patients hospitalised with COVID-19. Details of the trial design and results for other possible treatments (dexamethasone, hydroxychloroquine, lopinavir-ritonavir, azithromycin, tocilizumab, convalescent plasma, colchicine, aspirin, casirivimab plus imdevimab, and baricitinib) have been published previously.^6–8,13,18–23^ The trial is underway at 177 hospital organisations in the United Kingdom supported by the National Institute for Health and Care Research Clinical Research Network, and also at 15 non-UK hospitals (appendix pp 3-29). Of these, 27 UK hospitals participated in the DMF comparison. The trial is coordinated by the Nuffield Department of Population Health at the University of Oxford (Oxford, UK), the trial sponsor. The trial is conducted in accordance with the principles of the International Conference on Harmonisation–Good Clinical Practice guidelines and approved by the UK Medicines and Healthcare products Regulatory Agency (MHRA) and the Cambridge East Research Ethics Committee (ref: 20/EE/0101). The protocol and statistical analysis plan are included in the appendix (pp 61-172) with additional information available on the study website www.recoverytrial.net.

Patients admitted to hospital were eligible for the study if they had clinically suspected or laboratory confirmed COVID-19 and no medical history that might, in the opinion of the attending clinician, put the patient at significant risk if they were to participate in the trial. Those aged <18 years and pregnant women were not eligible for randomisation to DMF. Written informed consent was obtained from all patients, or a legal representative if patients were too unwell or otherwise unable to provide informed consent.

### Randomisation and masking

Baseline data were collected using a web-based case report form that included patient demographics, level of respiratory support, major comorbidities, suitability to receive the study treatment, and treatment availability at the study site (appendix pp 38-40). Eligible and consenting patients were assigned in a 1:1 ratio to either usual standard of care or usual standard of care plus DMF using web-based simple (unstratified) randomisation with allocation concealed until after randomisation (appendix pp 36-38). For some patients, DMF was unavailable at the hospital at the time of enrolment or was considered by the managing physician to be either definitely indicated or definitely contraindicated. These patients were not eligible for randomisation between DMF and usual care. Patients allocated DMF were to receive 120mg by mouth every 12 hours for the first 4 doses, followed by 240mg every 12 hours, for total treatment duration of 10 days or until hospital discharge, whichever was sooner. The stepped increase in dose was chosen to minimise flushing and gastrointestinal side effects, and the protocol also allowed dose reduction to a minimum of 120mg once daily if needed to control side effects.

As a platform trial, and in a factorial design, patients could be simultaneously randomised to other treatment groups: i) casirivimab plus imdevimab versus usual care, ii) aspirin versus usual care, iii) baricitinib versus usual care, and iv) empagliflozin versus usual care. Further details of when these factorial randomisations were open are provided in the supplementary appendix (pp 36-38). Participants and local study staff were not masked to the allocated treatment. The trial steering committee, investigators, and all other individuals involved in the trial were masked to outcome data during the trial.

### Procedures

Participants had daily assessment of clinical status from day 1 to day 10, using a seven-category ordinal scale as follows: 1) discharged alive; 2) in hospital, not requiring oxygen or medical care; 3) in hospital, not requiring oxygen but requiring medical care; 4) in hospital, requiring oxygen via simple face mask or nasal cannula; 5) in hospital, requiring high-flow nasal oxygen or non-invasive ventilation; 6) in hospital, requiring invasive mechanical ventilation or extracorporeal membrane oxygenation; and 7) dead.^24^ At baseline and on days 3, 5 and 10, the S/F_94_ ratio was recorded. The S/F_94_ ratio is defined as the ratio of peripheral oxygen saturations (SpO_2_) to the fraction of inspired oxygen (FiO_2_), with any supplemental oxygen reduced until SpO_2_ is <94% (patients were transferred to an oxygen delivery device providing a defined FiO_2_ if necessary). Details of S/F_94_ measurement and its rationale are outlined in the appendix (pp 30, 139-151). Derivation and evaluation of the S/F_94_ endpoint are reported in a companion paper.^25^ Blood C-reactive protein, creatinine and alanine or aspartate transaminase were measured on days 3, 5 and 10, along with treatment adherence and details of adverse events. The above details were collected into a web-based DMF follow up form developed for this early phase assessment, completed daily until day 10 (appendix pp 41-45). Another online follow-up form was completed when participants were discharged, had died or at 28 days after randomisation, whichever occurred earliest (appendix pp 46-53). This recorded information on receipt of other COVID-19 treatments, duration of admission, receipt of respiratory or renal support, and vital status (including cause of death). In addition, routine healthcare and registry data were obtained including information on vital status (with date and cause of death), discharge from hospital, receipt of respiratory support, or renal replacement therapy.

### Outcomes

The primary outcome was clinical status at day 5, as assessed on the ordinal scale. Secondary outcomes were: time to sustained improvement by at least one category on the ordinal scale from baseline (persisting for >1 day), time to discharge from hospital, S/F_94_ ratio at day 5, blood C-reactive protein at day 5, and improvement in clinical status by at least one category at day 10. The initial protocol specified day 5 S/F_94_ as the primary outcome and day 5 clinical status as a secondary outcome, but these were switched in October 2021 when it was realised that discharges before day 5 would lead to significant amounts of missing data for the S/F_94_ outcome. This decision was made by the trial investigators whilst blinded to the results of the DMF comparison.

Subsidiary clinical outcomes were: use of ventilation and, separately, use of renal dialysis or haemofiltration, among patients not on such treatment at randomisation, and thrombotic events. Pre-specified safety outcomes were: flushing, gastrointestinal symptoms, transaminitis (peak ALT/AST >3x upper limit of normal), acute kidney injury (peak creatinine >1.5x value at randomisation), cause-specific mortality, bleeding events, major cardiac arrhythmias, and non-coronavirus infections. Information on suspected serious adverse reactions was collected in an expedited fashion to comply with regulatory requirements.

### Statistical Analysis

The primary analysis for all outcomes was by intention-to-treat, comparing patients randomised to DMF with patients randomised to usual care. For the primary outcome of clinical status at day 5, the common odds ratio of a worse outcome with DMF versus usual care was estimated using ordinal logistic regression with adjustment for baseline score. For 20 participants still alive in hospital on day 5 without a recorded score, the median possible score was imputed. The proportional odds assumption was assessed and there was no evidence that this was violated (p-value from test of proportional odds assumption 0.95).

For time to sustained improvement, the log-rank observed minus expected statistic and its variance were used to test the null hypothesis of equal survival curves (i.e., the log-rank test) and to calculate the one-step estimate of the average rate ratio. Analyses were restricted to the first 10 days as ordinal scores were not collected after this. A similar analysis was used for time to discharge up to day 28, with patients who died in hospital right-censored on day 29. Median time to discharge was derived from Kaplan-Meier estimates.

Comparisons of S/F_94_ ratio and log-transformed CRP at day 5 were performed using ANCOVA adjusted for each participant’s baseline value. For patients who were discharged from hospital, for whom it was not possible to measure S/F_94_ ratio at day 5, a value of 4.76 was imputed (i.e. the maximum value, assuming saturations of 100% when breathing 21% oxygen). Multiple imputation methods were used to account for any other missing data.^26^ Risk ratios were used to compare treatment arms for improvement of clinical status at day 10, and for all subsidiary and safety outcomes.

Estimates of rate and risk ratios are shown with 95% confidence intervals. All p-values are 2-sided and are shown without adjustment for multiple testing. The full database is held by the study team, which collected the data from study sites and performed the analyses at the Nuffield Department of Population Health, University of Oxford (Oxford, UK).

It was estimated that enrollment of at least 700 patients would provide 80% power (at 2p=0.05) to detect a common odds ratio of 0.67, even if 10% of participants discontinued study treatment before day 5. Recruitment was halted on 19^th^ November 2021 after target recruitment had been reached. The Trial Steering Committee and all other individuals involved in the trial were masked to outcome data until 28 days after the close of recruitment.

Analyses were performed using SAS version 9.4 and R version 3.4. The trial is registered with ISRCTN (50189673) and clinicaltrials.gov (NCT04381936).

## RESULTS

Between 2 March 2021 and 18 November 2021, 713 (44%) of 1630 patients enrolled into the RECOVERY trial at sites participating in the DMF comparison were eligible to be randomly allocated to DMF (i.e. consent was obtained, DMF was available in the hospital at the time and the attending clinician was of the opinion that the patient had no known indication for or contraindication to DMF, figure 1). 356 patients were randomly allocated to DMF plus usual standard of care and 357 were randomly allocated to usual standard of care alone. The mean age of study participants in this comparison was 57.1 years (SD 15.7) and the median time since symptom onset was 9 days (IQR 7 to 11 days) (table 1). At randomisation, 40 (6%) patients did not require oxygen, 535 (75%) required simple oxygen without ventilation, and 135 (19%) required non-invasive ventilation. 674 (95%) were receiving corticosteroids.

**Figure 1:**
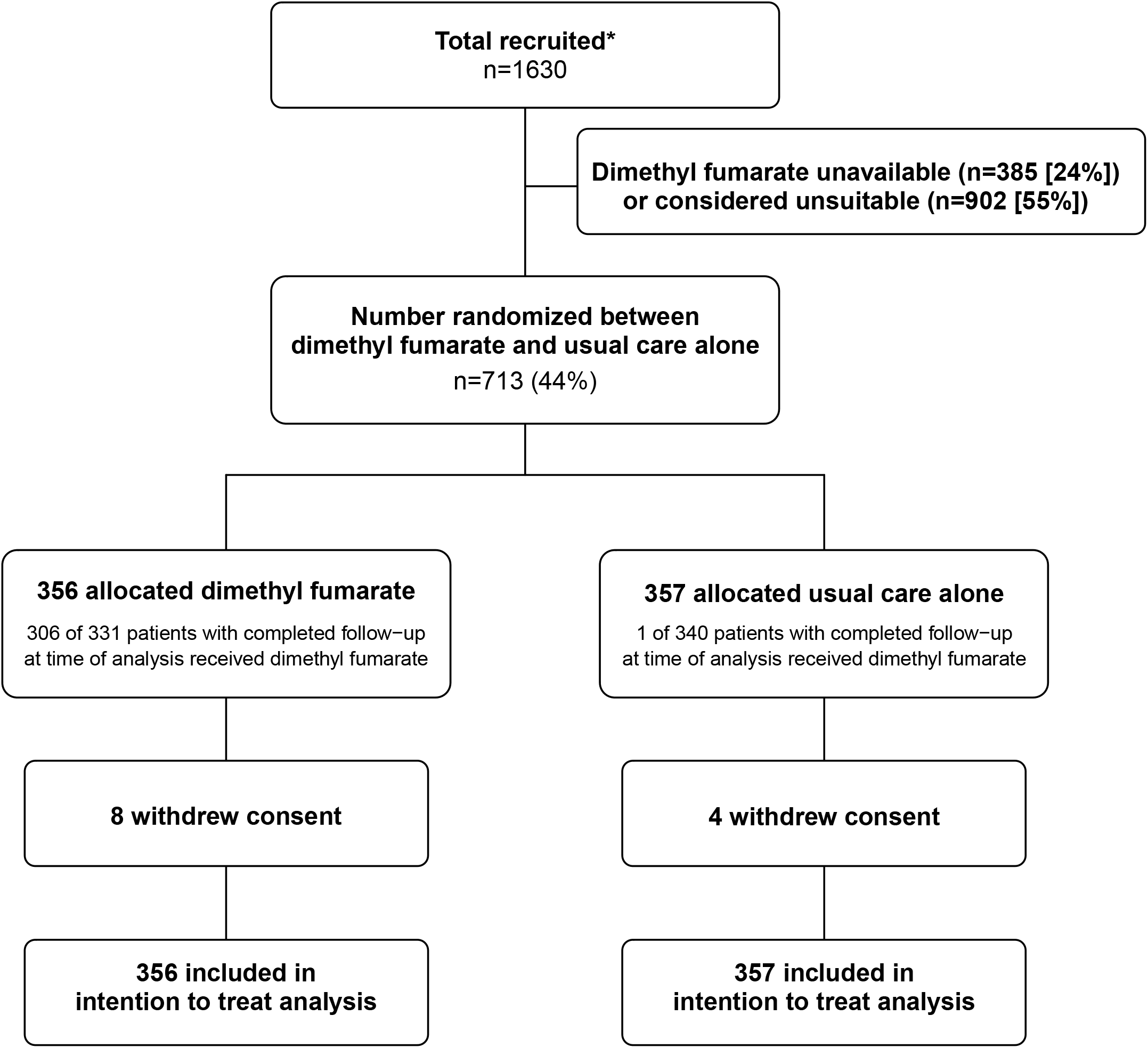
Trial profile. ITT=intention to treat. * Number recruited overall at sites participating in the DMF comparison during the period that adult participants could be recruited. DMF unavailable and DMF unsuitable are not mutually exclusive.

**Table 1:**
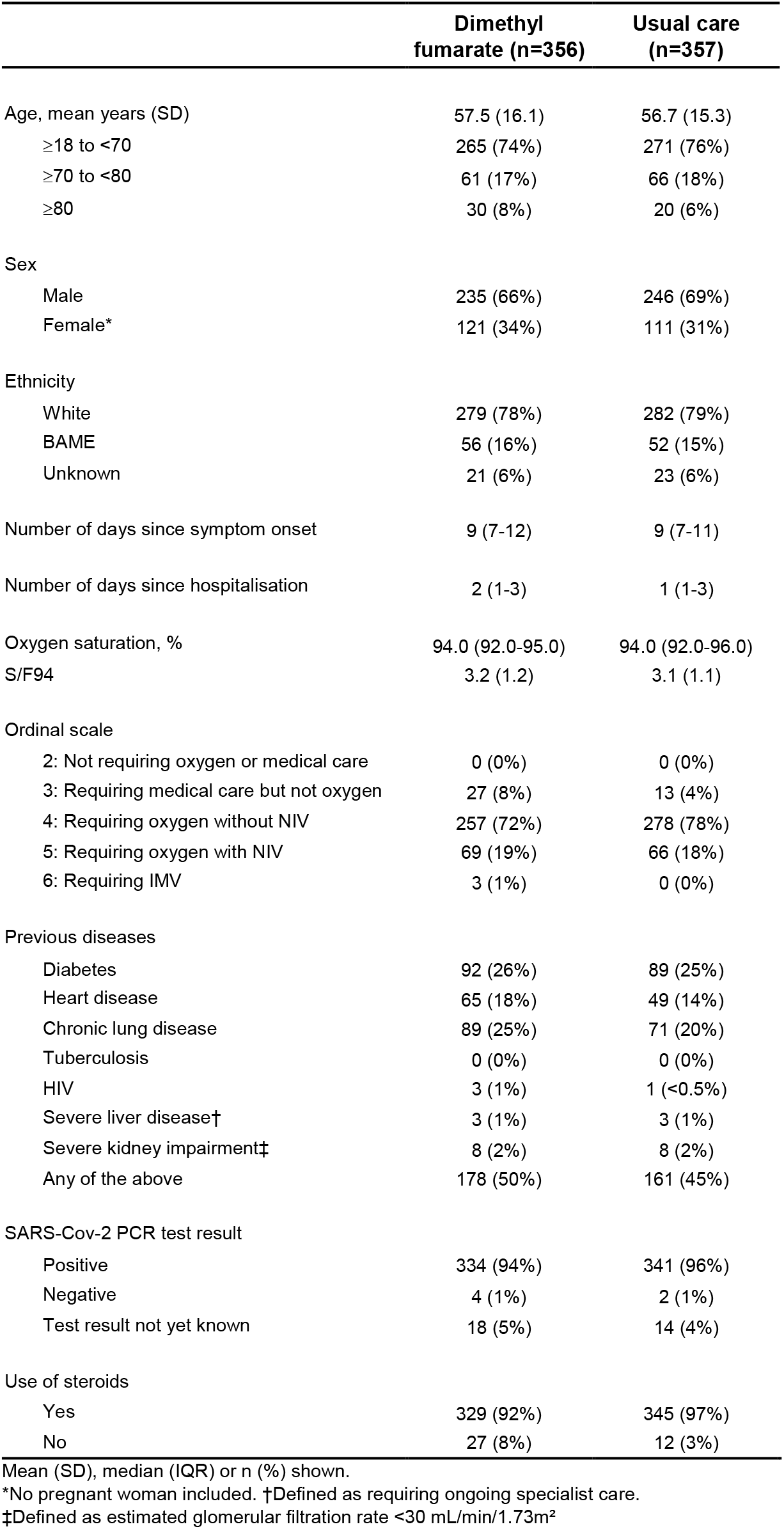
Baseline characteristics of patients randomised to dimethyl fumarate vs usual care.

Among patients with known DMF adherence, 306/331 (92%) allocated to DMF received at least one dose, and 248/331 (75%) received at least half of the specified treatment course. Use of other treatments for COVID-19 was similar among patients allocated DMF and those allocated usual care, including use of baricitinib (44% of participants), and tocilizumab or sarilumab (34% of participants) (webtable 1).

Primary outcome data are known for 693 (97%) of randomly assigned patients. There was no significant difference between the groups in clinical status at day 5 (common odds ratio of unfavourable outcome 1.12; 95% confidence interval [CI] 0.85–1.46; p=0.42; table 2, figures 2 and 3).

**Figure 2:**
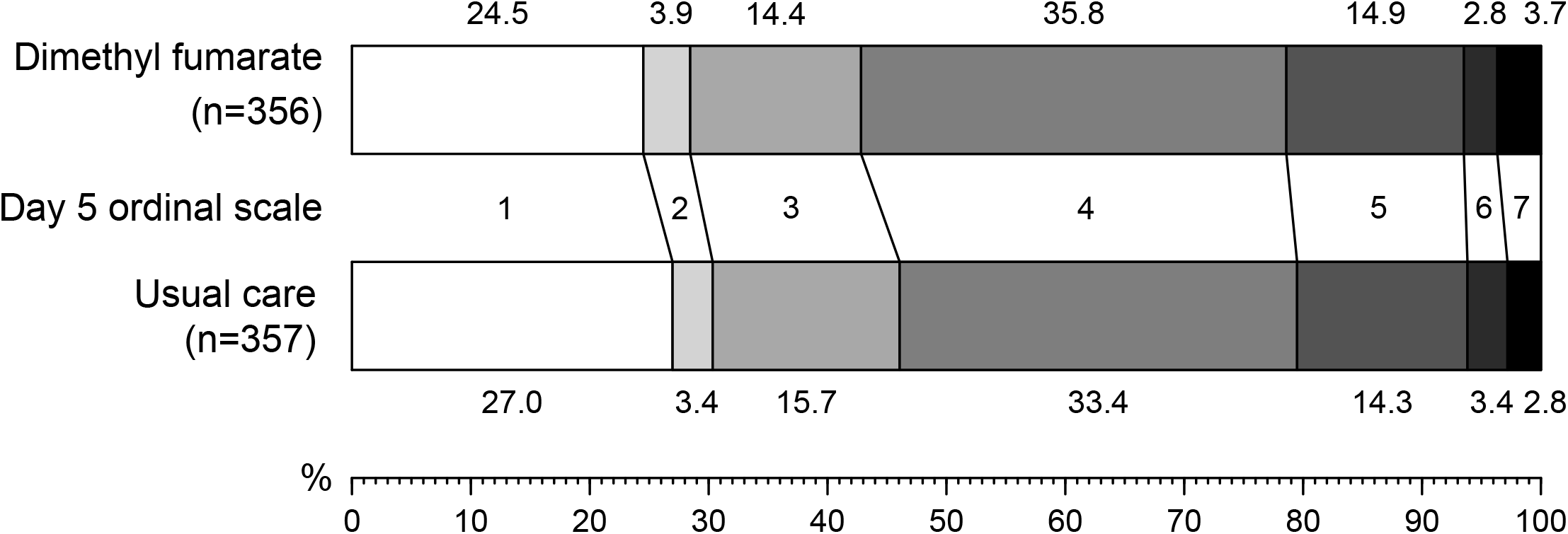
Distribution of clinical ordinal scale at 5 days by randomised allocation.

**Figure 3:**
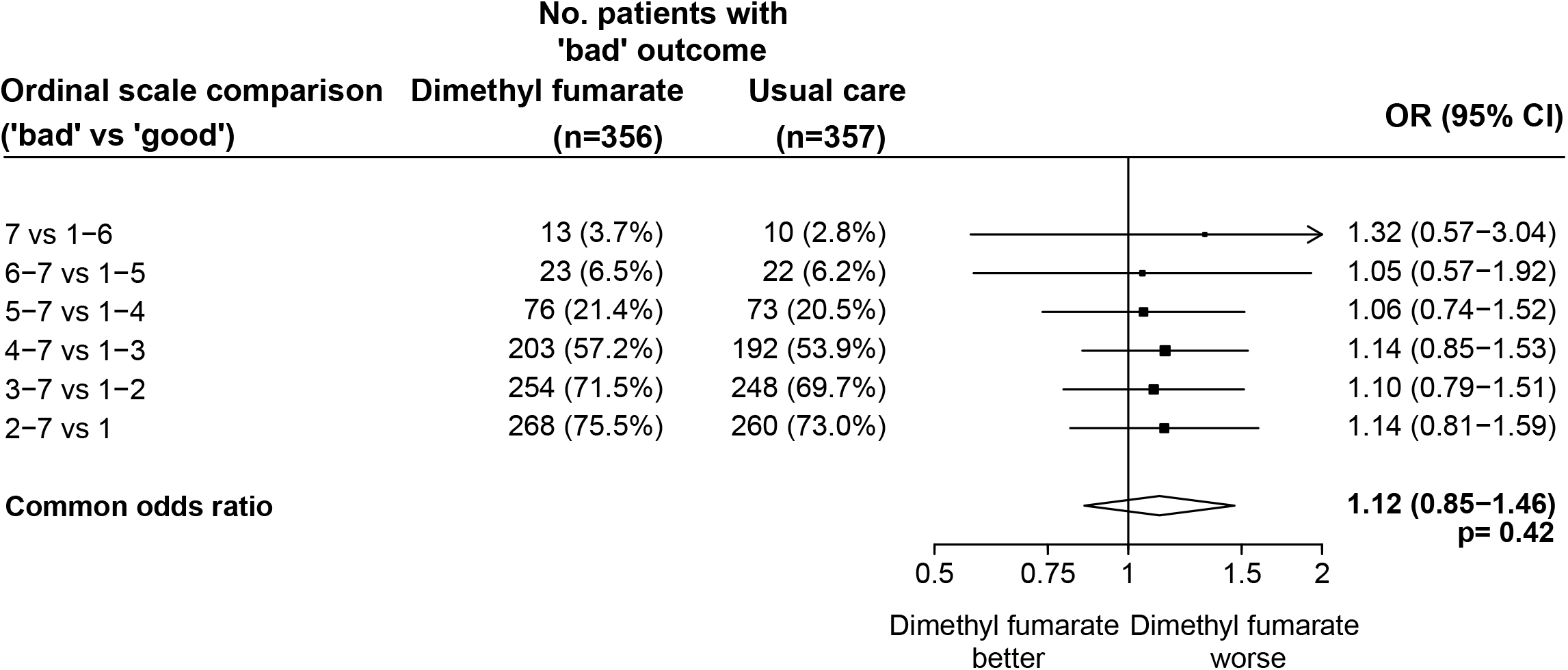
Effects of allocation to dimethyl fumarate on relative odds of a bad outcome on the clinical ordinal scale at day 5, for each alternative definition of bad outcome. Odds ratio estimates for each ordinal scale comparison are represented by squares (with areas of the squares proportional to the amount of statistical information) and the lines through them correspond to the 95% CIs.

**Table 2:** Effect of allocation to dimethyl fumarate on key study outcomes.

|  | Dimethyl fumarate<br>(n=356) | Usual care (n=357) | Treatment effect<br>(95% CI) | p value |
| --- | --- | --- | --- | --- |
| **Primary outcome** |  |  |  |  |
| Ordinal scale at day 5* |  |  |  |  |
| 7 vs 1-6 | 13 (3.7%) | 10 (2.8%) |  |  |
| 6-7 vs 1-5 | 23 (6.5%) | 22 (6.2%) |  |  |
| 5-7 vs 1-4 | 76 (21.4%) | 73 (20.5%) |  |  |
| 4-7 vs 1-3 | 203 (57.2%) | 192 (53.9%) |  |  |
| 3-7 vs 1-2 | 254 (71.5%) | 248 (69.7%) |  |  |
| 2-7 vs 1 | 268 (75.5%) | 260 (73.0%) |  |  |
| Common odds ratio |  |  | 1.12 (0.85-1.46) | 0.42 |
| **Secondary clinical outcomes** |  |  |  |  |
| Sustained improvement in ordinal category within 10 days† | 246 (69.1%) | 258 (72.3%) | 0.96 (0.80-1.16) | 0.70 |
| Improvement in clinical status at day 10§ | 246 (69%) | 259 (73%) | 0.95 (0.87-1.05) | 0.31 |
| Baseline-adjusted day 5 S/F94‡ | 3.58 (0.06) | 3.64 (0.06) | -0.06 (-0.22 to 0.10) | 0.45 |
| Baseline-adjusted day 5 CRP§ | 14.4 (1.2) | 14.0 (1.2) | 2% (-18 to 29%) | 0.84 |
| Median duration of hospitalization, days | 8 | 8 |  |  |
| Discharged from hospital alive within 28 days† | 274 (77.0%) | 281 (78.7%) | 0.95 (0.80-1.13) | 0.59 |
| **Subsidiary clinical outcomes** |  |  |  |  |
| Use of ventilation\$\|\| | 58/284 (20%) | 60/291 (21%) | 0.99 (0.72-1.37) | 0.95 |
| Non-invasive ventilation | 56/284 (20%) | 56/291 (19%) | 1.02 (0.73-1.43) | 0.89 |
| Invasive mechanical ventilation | 14/284 (5%) | 12/291 (4%) | 1.20 (0.56-2.54) | 0.64 |
| Successful cessation of invasive mechanical ventilation** | 0/3 (0.0%) | 0/0 | - | - |
| Renal dialysis or haemofiltration\$\|\| | 7/356 (2%) | 6/355 (2%) | 1.16 (0.39-3.43) | 0.78 |
| **Safety outcomes** |  |  |  |  |
| Flushing\$ | | | | |
| Some | 23 (7%) | 11 (3%) | 2.08 (1.03-4.21) | 0.04 |
| Severe | 8 (2%) | 0 (0%) | - | - |
| Subtotal: Any flushing | 31 (9%) | 11 (3%) | 2.81 (1.44-5.50) | 0.0026 |
| Gastrointestinal symptoms\$ | | | | |
| Some | 34 (10%) | 18 (5%) | 1.88 (1.08-3.27) | 0.02 |
| Severe | 4 (1%) | 1 (0%) | 3.99 (0.45-35.51) | 0.21 |
| Subtotal: Any gastrointestinal symptoms | 38 (11%) | 19 (5%) | 1.99 (1.17-3.39) | 0.01 |
| Transaminitis\$ | 57 (19%) | 56 (18%) | 1.05 (0.75-1.46) | 0.78 |
| Acute kidney injury\$ | 9 (3%) | 12 (4%) | 0.75 (0.32-1.75) | 0.51 |
| Non-coronavirus infection |  |  |  |  |
| Pneumonia | 18 (5%) | 20 (6%) | 0.90 (0.49-1.68) | 0.75 |
| Urinary tract | 1 (0%) | 4 (1%) | 0.25 (0.03-2.23) | 0.21 |
| Biliary | 0 (0.00%) | 0 (0.00%) | - | - |
| Other intra-abdominal | 1 (0.28%) | 1 (0.28%) | - | - |
| Blood stream | 4 (1%) | 1 (0%) | 4.01 (0.45-35.71) | 0.21 |
| Skin | 1 (0.28%) | 1 (0.28%) | - | - |
| Other | 6 (2%) | 6 (2%) | 1.00 (0.33-3.08) | 1.00 |
| Subtotal: Any non-coronavirus infection | 27 (8%) | 31 (9%) | 0.87 (0.53-1.43) | 0.59 |

\*Number of patients with a 'bad' outcome given. Treatment effects are odds ratios for 'bad' vs 'good' outcome. Common odds ratio estimated using a proportional odds model adjusted for ordinal scale at randomisation. For the 20 patients with missing data on ordinal scale at day 5, the median possible category was imputed (rounded up when there are an even number of possibilities).
†Treatment effect is a rate ratio estimated using logrank methods.
‡Treatment effect is difference in mean S/F94 estimated using ANCOVA with adjustment for baseline S/F94. For patients discharged alive by day 5, a value of 4.76 was imputed. All 135 (18.9%) other missing values at day 5 were imputed using multiple imputation.
§Treatment effect is a risk ratio.
§ANCOVA analyses of log transformed CRP with adjustment for randomisation value were conducted. 276 (38.7%) missing values at day 5 imputed using multiple imputation. Geometric means and approximate standard errors are presented and treatment effect is percentage change in CRP.
||Analyses include only those on no ventilation support at randomisation.
\*\*Analyses restricted to those on invasive mechanical ventilation at randomisation.
||Analyses exclude those on haemodialysis or haemofiltration at randomisation.

We found no evidence of an effect of DMF on any secondary or subsidiary outcome (table 2). There was no significant difference in the time to sustained clinical improvement (rate ratio 0.96; 95% CI 0.80–1.16, p=0.70) or time to discharge from hospital alive (rate ratio 0.95, 95% CI 0.80–1.13, p=0.59). At day 5 after randomisation there was no significant difference in S/F_94_ (difference in mean S/F_94_ -0.06; 95% CI -0.22 to 0.10; p=0.45) or in CRP (difference in geometric mean 2%; 95% CI -18% to 29%; p=0.84). The proportion of patients with improvement of clinical status by day 10 was similar in both groups (risk ratio 0.95; 95% CI 0.87–1.05; p=0.31).

Compared to usual care, more participants allocated to DMF suffered flushing (9% vs 3%, risk ratio 2.81; 95% CI 1.44–5.50; p=0.003) and gastrointestinal symptoms (11% vs 5%, risk ratio 1.99; 95% CI 1.17–3.39; p=0.01, table 2). DMF treatment was discontinued because of adverse events in 42 (13%) patients, mainly because of flushing, rash, diarrhoea, or abnormal liver function tests, and 12 (4%) patients required DMF dose reduction (webtable 2). A further 32 (10%) patients discontinued DMF for reasons other than adverse events, mainly because they were no longer able to take tablets (webtable 2). There was one report of a serious adverse reaction believed related to DMF, in a patient whose ALT rose to 5 times the upper limit of normal, although the total number of patients with transaminitis reported was similar in both groups (19% vs 18%, risk ratio 1.05; 95% CI 0.75-1.46; p=0.78, table 2). There was no evidence of an effect of DMF on other safety outcomes, including all-cause mortality, cause-specific mortality, cardiac arrhythmia, non-coronavirus infections, acute kidney injury, thrombotic events or bleeding events (table 2, webtables 3-5).

## DISCUSSION

In this initial evaluation in the RECOVERY trial, involving over 700 patients hospitalised with COVID-19, treatment with DMF was not associated with improvement in any clinical outcome compared with usual care alone. This is the first randomised trial of DMF for the treatment of COVID-19, and although pre-clinical data suggest that it interferes with inflammatory pathways important to the pathogenesis of COVID-19 pneumonia, this did not translate into any evident benefit of treatment.

Inflammasome-mediated inflammation is activated in patients with severe COVID-19, making it a promising therapeutic target. DMF effectively inhibits inflammasome activation in vitro and is effective as an anti-inflammatory treatment for psoriasis and relapsing-remitting multiple sclerosis (where it halves the rate of relapse).^15–17^ However, colchicine and DMF have both now been evaluated in hospitalised COVID-19 patients because they interfere with inflammasome activation, and neither has produced any discernible improvement in outcome. This may be because these agents do not block this pathway effectively enough, or because activation of this pathway is not causally related to disease trajectory, at least among hospitalised patients receiving current standard treatment. Corticosteroids were received by 95% of the trial population, and a significant proportion also received an IL-6 inhibitor or JAK inhibitor. It is possible that DMF could have had a beneficial effect in the absence of other immunomodulators, but it appears to add little or nothing to current usual care.

Treatment was discontinued because of adverse events in 13% of patients, mostly because of flushing, rash, and gastrointestinal side-effects. These are recognised side-effects of DMF, although rarely caused discontinuation in outpatient placebo-controlled trials in patients with multiple sclerosis.^16,17^ Other than these adverse effects, no safety concerns of DMF treatment were identified. DMF was discontinued because of abnormal liver function tests in 6 patients, but ALT elevations are commonly seen in hospitalised patients with COVID and occurred in 18% of participants in the usual care arm.^27^ The proportion of patients with transaminitis was similar in the DMF and usual care groups, suggesting DMF was not a significant cause of transaminitis, and highlighting the need for systematic data collection when evaluating adverse events in an open label study.

Strengths of this trial include that it was randomised, had broad eligibility criteria, and follow up was 97% complete. However, there are some limitations: as an early phase study, it was not large enough to rule out a benefit in mortality, nor to assess whether treatment effects might have varied among specific groups of patients. The trial was open label, so participants and local hospital staff were aware of the assigned treatment. This could potentially affect the assessment of some outcomes, particularly if allocation to DMF led to patients staying in hospital to receive treatment rather than being discharged. However, our protocol specified that treatment was to stop when patients were ready for discharge, and the distribution of clinical status at day 5 provides no evidence to suggest that otherwise healthy patients stayed in hospital to receive DMF (Figures 2 and 3). Finally, we only studied patients who had been hospitalised with COVID-19, so do not provide any evidence on the safety and efficacy of DMF in other groups, such as outpatients.

In summary, the results of this randomised trial do not support the use or further study of DMF in adults hospitalised with COVID-19.

### Evidence before this study

We searched PubMed, medRxiv, bioRxiv, and the WHO International Clinical Trials Registry Platform from September 1, 2019 to July 31, 2022 for clinical trials evaluating the effect of dimethyl fumarate treatment in patients with COVID-19, using the search terms (SARS-CoV-2 OR COVID OR COVID-19 OR 2019-nCoV OR Coronavirus) AND (dimethyl fumarate OR Tecfidera OR Skilarence OR BG-12). We did not identify any reported trials.

### Added value of this study

The Randomised Evaluation of COVID-19 therapy (RECOVERY) trial is the first randomised trial to report results of the effect of dimethyl fumarate in patients with COVID-19. We found no significant effect of DMF compared with usual care alone on clinical status at day 5, or any other clinical outcomes.

### Implications of all the available evidence

There is no evidence that treatment with dimethyl fumarate is of clinical benefit for adults hospitalised with COVID-19 compared with current usual care.

### Contributors

This manuscript was initially drafted by LP, PWH and MJL. All authors contributed to data interpretation and critical review and revision of the manuscript. PWH and MJL vouch for the data and analyses, and for the fidelity of this report to the study protocol and data analysis plan. PWH, MM, JKB, MHB, JD, SNF, TJ, KJ, EJ, MK, WSL, AMo, AMu, KR, GT, RH, and MJL designed the trial and study protocol. LP, MC, G P-A, MM, RS, BP, AU, CAG, DJD, FM, JM, PC, JS, BY, and the Data Linkage team at the RECOVERY Coordinating Centre, and the Health Records and Local Clinical Centre staff listed in the appendix collected the data. NS and JRE had access to the study data and did the statistical analysis. PWH and MJL had final responsibility for the decision to submit for publication.

### Writing Committee (on behalf of the RECOVERY Collaborative Group)

Peter W Horby,* Leon Peto,* Natalie Staplin,* Mark Campbell, Guilherme Pessoa-Amorim, Marion Mafham, Jonathan R Emberson, Richard Stewart, Benjamin Prudon, Alison Uriel, Christopher A Green, Devesh J Dhasmana, Flora Malein, Jaydip Majumdar, Paul Collini, Jack Shurmer, Bryan Yates, J Kenneth Baillie, Maya H Buch, Jeremy Day, Saul N Faust, Thomas Jaki, Katie Jeffery, Edmund Juszczak, Marian Knight, Wei Shen Lim, Alan Montgomery, Andrew Mumford, Kathryn Rowan, Guy Thwaites, Richard Haynes,^†^ Martin J Landray.^†^

### Data Monitoring Committee

Peter Sandercock, Janet Darbyshire, David DeMets, Robert Fowler, David Lalloo, Mohammed Munavvar, Adilia Warris, Janet Wittes.

## Supporting information

CONSORT checklist

## Data Availability

All data produced in the present study are available upon reasonable request to the authors

## Data sharing

The protocol, consent form, statistical analysis plan, definition & derivation of clinical characteristics & outcomes, training materials, regulatory documents, and other relevant study materials are available online at www.recoverytrial.net. As described in the protocol, the trial Steering Committee will facilitate the use of the study data and approval will not be unreasonably withheld. Deidentified participant data will be made available to bona fide researchers registered with an appropriate institution within 3 months of publication. However, the Steering Committee will need to be satisfied that any proposed publication is of high quality, honours the commitments made to the study participants in the consent documentation and ethical approvals, and is compliant with relevant legal and regulatory requirements (e.g. relating to data protection and privacy). The Steering Committee will have the right to review and comment on any draft manuscripts prior to publication. Data will be made available in line with the policy and procedures described at: https://www.ndph.ox.ac.uk/data-access. Those wishing to request access should complete the form at https://www.ndph.ox.ac.uk/files/about/data_access_enquiry_form_13_6_2019.docx and e-mail to

## Acknowledgements

Above all, we would like to thank the patients who participated in this trial. We would also like to thank the many doctors, nurses, pharmacists, other allied health professionals, and research administrators at NHS hospital organisations across the whole of the UK, supported by staff at the National Institute of Health Research (NIHR) Clinical Research Network, NHS DigiTrials, Public Health England, Department of Health & Social Care, the Intensive Care National Audit & Research Centre, Public Health Scotland, National Records Service of Scotland, the Secure Anonymised Information Linkage (SAIL) at University of Swansea, and the NHS in England, Scotland, Wales and Northern Ireland.

The RECOVERY trial is supported by grants to the University of Oxford from UK Research and Innovation (UKRI) and NIHR (MC_PC_19056), the Wellcome Trust (Grant Ref: 222406/Z/20/Z) through the COVID-19 Therapeutics Accelerator, and by core funding provided by the NIHR Oxford Biomedical Research Centre, the Wellcome Trust, the Bill and Melinda Gates Foundation, the Foreign, Commonwealth and Development Office, Health Data Research UK, the Medical Research Council Population Health Research Unit, the NIHR Health Protection Unit in Emerging and Zoonotic Infections, and NIHR Clinical Trials Unit Support Funding. TJ is supported by a grant from UK Medical Research Council (MC_UU_0002/14). WSL is supported by core funding provided by NIHR Nottingham Biomedical Research Centre. Tocilizumab was provided free of charge for this trial by Roche Products Limited. Regeneron Pharmaceuticals supported the trial through provision of casirivimab and imdevimab. The views expressed in this publication are those of the authors and not necessarily those of the NHS, the NIHR, or the UK Department of Health and Social Care. For the purpose of Open Access, the author has applied a CC BY public copyright licence to any Author Accepted Manuscript version arising from this submission.

